# Budgeting and advocacy to improve water, sanitation, and hygiene in healthcare facilities: a case study in Nepal

**DOI:** 10.1101/2024.01.29.24301941

**Authors:** Laxman Kharal Chettry, Prakash Bohara, Ramesh C. Bohara, Ram Hari Jajal, Sarad Khadha, Hari Subedi, Debesh Giri, Sarbesh Sharma, Upendra Dhungana, Matteus Thijs van der Valen, John Brogan, Darcy M. Anderson

**Affiliations:** Terre des hommes (Tdh) Foundation, Nepal Country Office, Lalitpur, Nepal; Swiss Water and Sanitation Consortium, Zurich, Switzerland; Thakurbaba Municipality, Bardiya District, Lumbini Province, Nepal; Geruwa Rural Awareness Association, Gulariya Municipality-5, Bardiya, Nepal; Management Division, Department of Health Services / Ministry of Health and Population, Kathmandu, Nepal; Helvetas, Zurich, Switzerland; The Water Institute at UNC, Gillings School of Global Public Health, the University of North Carolina at Chapel Hill, Chapel Hill, NC, USA

## Abstract

Barriers to achieving and sustaining access to water, sanitation, hygiene, waste management (collectively, “WASH”) in healthcare facilities include a supportive policy environment and adequate funding. While guidelines exist for assessing needs and making initial infrastructure improvements, there is little guidance on how to develop budgets and policies to sustain WASH services in the long-term. We conducted costing and advocacy activities in Thakurbaba municipality, Nepal, with the aim of developing a budget and operations and maintenance policy for WASH in healthcare facilities in partnership with the municipal government. Our objectives for this study are to (1) describe the process and methods used for costing and advocacy, (2) report the costs to achieve and maintain basic WASH services in the eight healthcare facilities of Thakurbaba municipality, and (3) report the outcomes of advocacy activities and policy development. We applied bottom-up costing to enumerate the resources necessary to achieve and maintain basic WASH services and their costs. The annual costs of WASH services ranged from USD 4,881 to 9,527 (including operations and maintenance and annualized capital investments). Cost findings were used to prepare annual budgets recommended to achieve and maintain basic access, which were presented to municipal government and incorporated into an operations and maintenance policy. To-date, the municipality has adopted the policy and established a recovery fund of USD 3,831 for repair and maintenance of infrastructure, and an additional USD 192 per facility for discretionary WASH spending. Advocacy at the national level for WASH in healthcare facilities is currently being championed by the municipality, and findings from this project are being used to inform development of a nationally costed plan for universal access. This study is intended to provide a roadmap for how cost data can be collected and applied to inform policy.

## Introduction

Environmental conditions for water, sanitation, hygiene, cleaning, and waste management (hereafter collectively called “WASH”) in healthcare facilities are critical for safe care delivery and a well-functioning health system. In recognition of their importance, the World Health Organization (WHO) and United Nations Children’s Fund (UNICEF) have published guidelines for^1^ eight recommended steps for countries to achieve universal coverage.^2^ These guidelines— often called the “Eight Practical Steps”—have been widely adopted as a framework for guiding national action, and the WHO and UNICEF currently track progress on these steps for over 70 countries.^3^ The first steps are conducting situation assessments, setting targets, and developing national costed roadmaps for achieving those targets, which are considered key preparation before widespread program implementation.

Nepal has begun the Eight Practical Steps, including situation assessments and settings targets. In 2021, data from the WHO and UNICEF showed that 94% of healthcare facilities had an improved water source, 89% had improved sanitation, 97% had hand hygiene facilities at points of care, and 1% followed waste management procedures for safe segregation, treatment, and disposal.^4^ In 2018, the government of Nepal released draft national standards for WASH in healthcare facilities,^5^ which were broadly aligned with indicators recommended and used to measure basic and advanced access under the Joint Monitoring Program (JMP) of the WHO and UNICEF.^1,6^ The government formally endorsed these standards in 2021. Subsequent steps for developing a costed roadmap for achieving these standards are in-progress; a draft exists but is not yet finalized.

Funding for WASH in healthcare facilities in Nepal is disbursed from the federal government to the provincial level, which is subsequently disbursed to municipalities. Some funding is earmarked for specific major infrastructure projects (e.g., construction of new healthcare facilities). However, a substantial portion of funding is for discretionary spending, and municipal governments have broad authority to allocate this funding based on local needs and priorities.^7^ Under Nepal’s federal government system and relatively recent constitution—adopted in 2015— municipal governments also hold considerable authority for policy making. The Local Governance Operation Act of 2017 gives municipalities the authority to adopt acts, regulations, and working procedures per their specific needs, including WASH service delivery.^8^ However, technical expertise among municipal governments for planning and budgeting is often low, and non-governmental organizations (NGOs) play an important role in providing technical support and evidence to inform decision making.^9–11^

In 2022, the NGO Terre des hommes Nepal conducted costing and advocacy activities in Thakurbaba municipality, with the aim of accompanying the municipality to create budgets and an operations and maintenance (O&M) policy for WASH in healthcare facilities. To-date, these activities have successfully generated budgets for annual operating costs, which have been integrated into a policy for O&M. The municipal government has formally adopted the policy, allocated funding for its implementation, and begun deploying those funds to improve O&M. These outcomes are a meaningful achievement for municipal governments under the relatively new federalist government system and an important step for capacity building and systems strengthening. Cost estimates and learnings from these activities have informed national-level advocacy and contributed to ongoing efforts to develop nationally costed roadmaps for WASH in healthcare facilities. In this study, we report on these activities. This paper is intended to provide a case study for how costs data can inform advocacy and policy making for WASH in healthcare facilities, particularly for executing the Eight Practical Steps.

Our study objectives were to (1) describe the process and methods used for costing and advocacy, (2) assess the costs of achieving and sustaining basic WASH services in the eight healthcare facilities of Thakurbaba municipality, and (3) report the outcomes of policy development and advocacy activities. In the methods, we describe the key activities for the data collection and advocacy. In the results, we report the data from costing and the outcomes from policy development and advocacy activities conducted to-date. In the discussion, we reflect on lessons learned throughout this process.

## Methods

### Overview of data collection and advocacy activities

Costing, policy development, and advocacy activities were carried out in stages: (1) Tool development, (2) Initiation workshop, (3) Preliminary costing, (4) Sharing and discussion workshop, (5) Data validation and certification, (6) Budget calculations, (7) O&M Policy finalization and approval, (8): Dissemination and advocacy.

These activities were focused on achieving and maintaining basic WASH services. We defined basic service as meeting the basic indicators outlined by the WHO and UNICEF under the JMP.^1^ Healthcare facilities also included a small number of additional items not included under JMP indicators for basic service (e.g., fencing) or that went beyond basic to advanced service levels (e.g., water quality testing and treatment, additional toilets and hand hygiene facilities) that they considered necessary to provide adequate WASH. Supplemental files 1-8 indicate all included line items.

Our final list of WASH services included in costing and policy development were: water (source and pump, pipe network, water tower or storage system, treatment), sanitation (toilets and associated hand hygiene facilities, septic or other containment systems), hygiene (hand hygiene facilities at points of care), environmental cleaning, and waste management (sluice room or other waste storage and processing area, autoclave, waste pit, placenta pit, drainage), and fencing.

### Setting

This study was conducted in Thakurbaba municipality, in the Lumbini province of western Nepal. Thakurbaba municipality contains ten healthcare facilities, one in each of its nine wards and the Neulapur Municipal Hospital. We excluded two from this study: the Neulapur hospital, which was under construction at the time of this research, and the Godana Basic Health Center because it is located in the government forestry office and does not have its own WASH infrastructure. Demographic information on the eight included healthcare facilities is provided in Table 1.

**Table 1.**
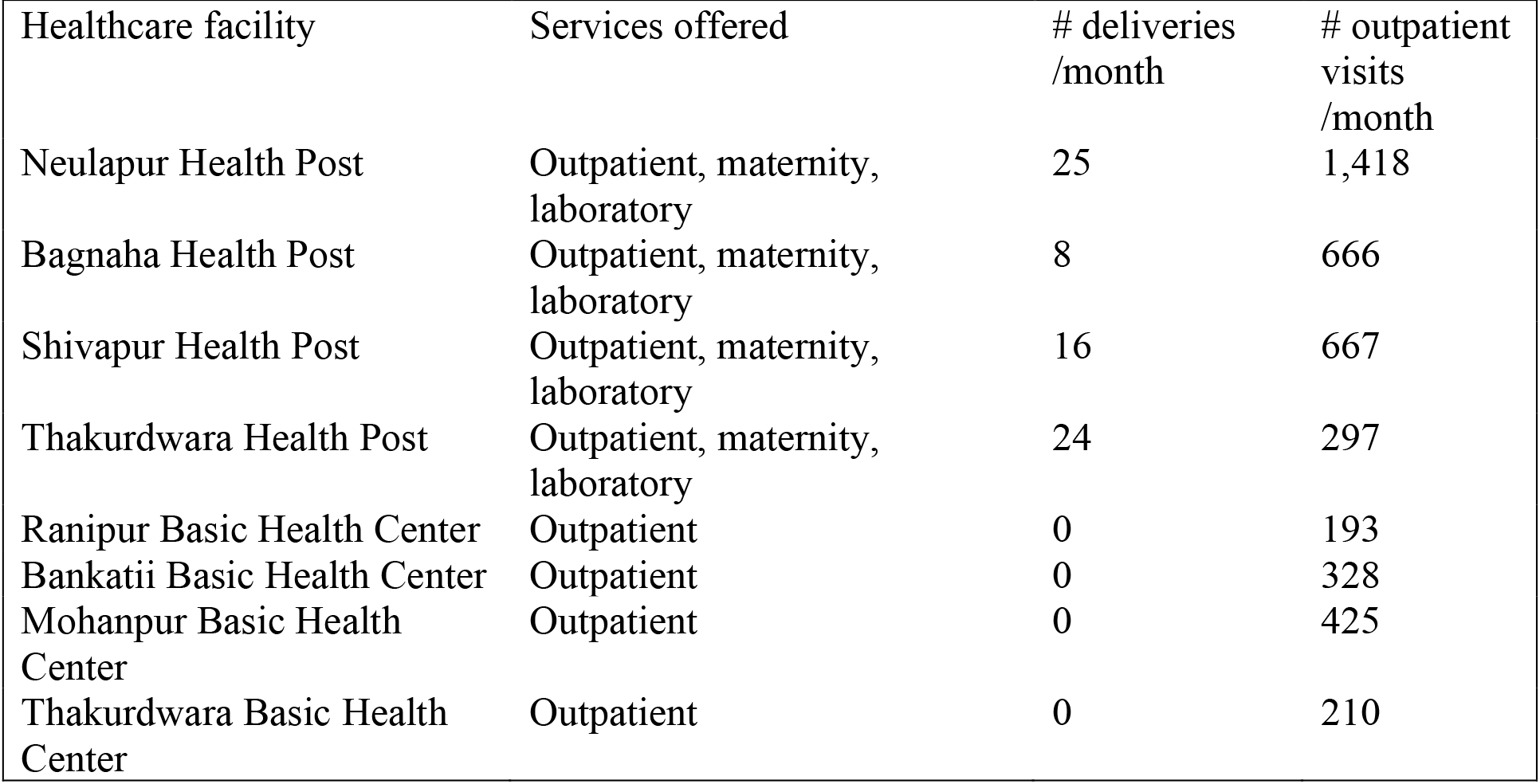
Demographics of healthcare facilities included in the study sample.

| Healthcare facility | Services offered | # deliveries /month | # outpatient visits /month |
| --- | --- | --- | --- |
| Neulapur Health Post | Outpatient, maternity, laboratory | 25 | 1,418 |
| Bagnaha Health Post | Outpatient, maternity, laboratory | 8 | 666 |
| Shivapur Health Post | Outpatient, maternity, laboratory | 16 | 667 |
| Thakurdwara Health Post | Outpatient, maternity, laboratory | 24 | 297 |
| Ranipur Basic Health Center | Outpatient | 0 | 193 |
| Bankatii Basic Health Center | Outpatient | 0 | 328 |
| Mohanpur Basic Health Center | Outpatient | 0 | 425 |
| Thakurdwara Basic Health Center | Outpatient | 0 | 210 |

#### Stage 1 Tool development

We created a costing tool in Microsoft Excel, based on previously developed methods.^12–14^ We chose Excel in part because project personnel were familiar with it, and the tabular format was well suited to data entry. We used bottom-up costing, in which all resources used in WASH provision are enumerated, a quantity and unit price are estimated for each resource, and total costs are calculated based on quantities and unit price. We pre-populated the spreadsheet with resources essential for achieving basic WASH, based on previous field research.^14^ We conducted a pilot visit to one healthcare facility, where we observed conditions and availability of infrastructure, goods, and services related to WASH. We also conducted a meeting to understand how repair tasks were carried out. This information was used to refine the list of resources pre-populated in the spreadsheet.

The final spreadsheet contained costs for initial capital investments (hardware and software) and annual O&M (maintenance, personnel, recurrent training, consumables). Definitions and examples for each cost type are provided in Table 2.

**Table 2.** Categories of expenses included in costing. WASH = water, sanitation, hygiene, cleaning, and waste management.

| Cost category | Definition | Example included expenses |
| --- | --- | --- |
| Capital hardware | Infrastructure or equipment purchases required to establish WASH services or implement changes to service delivery method that are not consumed during normal service operation | Sanitation facilities (superstructure with squat pan/seats, pit/septic tank)<br>Water source and pipe network |
| Capital software | Planning, procurement, and/or initial training costs associated with establishing | Initial infection prevention and operations and maintenance |
|  | new WASH services or implementing changes to WASH service delivery method | trainings delivered upon establishing infrastructure |
| Maintenance | Expenses required to repair and maintain functionality of capital hardware, including labor costs required for these purposes | Breakdown repairs (e.g., clogged pipes)<br>Cleaning of toilets, patient care areas<br>Supplies for water system testing (e.g., arsenic, residual chlorine) |
| Recurrent software | Necessary trainings, behavior change, and other non-tangible produced to be delivered each year for the upkeep of the established and other introduced practices. | Annual infection prevention training<br>Annual WASH FIT meetings |
| Personnel | Labor costs associated with normal operation of a service, including staff benefits; labor costs for maintenance (e.g., plumbers and repair technicians that are outsourced) are included under maintenance | WASH focal person<br>Support staff |
| Consumables | Products and supplies that are consumed during normal operation | Handwashing soap<br>Cleaning detergents<br>Cleaning tools (mops, brooms) |
| Support | Expenses required to strengthen WASH provision but that do not have direct service outputs | Communication<br>Capacity building |

#### Stage 2 Initiation workshop

We held an initiation workshop with approximately 40 key stakeholders, who were elected officials from municipal government (e.g., mayor, deputy mayor), bureaucrats and technical experts (e.g., WASH engineers, information technology specialists), representatives from NGOs working locally on WASH, leaders from healthcare facilities that would be included in the project, and the media. During the workshop, we explained that the purpose of the project was to develop an O&M policy for WASH in healthcare facilities and that costing would be done to develop budgets for the policy and provide a basis for evidence-based annual resource allocations.

The municipality formed a policy formulation committee, with the vice mayor as the formal chair, following their standard committee format. This committee was formed to steer the policy drafting process. A task force group comprised of three members—a local NGO representative participating in the project, the municipality health coordinator, and the municipality information technology officer—was formed to support the policy formulation committee. The task force supported tasks such as collecting and reviewing information and organizing consultations. Cost data collection and budgeting activities were conducted by the international NGO Terre des hommes and the local NGO Geruwa in collaboration with the Thakurbaba Municipality. The Water Institute at the University of North Carolina at Chapel Hill provided technical support.

#### Stage 3 Preliminary costing

We met with each healthcare facility; participants included the HCF in-charge, storekeeper, and nursing and support staff. During the meeting, the data collection team and meeting participants listed the type (e.g., pit latrine vs. pour-flush latrine) and quantity of all WASH infrastructure available at the healthcare facility. For each piece of infrastructure, healthcare facility staff were asked to describe its functionality in terms of number of breakdowns per year and average duration of breakdown. We cross-referenced this list of infrastructure with our prepopulated costing spreadsheet and revised as necessary. An engineer then estimated the costs for installation for all infrastructure (capital hardware and software), based on recall, the costs of similar items being constructed, and records that were readily available (e.g., regularly purchased items). Healthcare facility staff estimated the specific products, quantities, and costs for maintenance, recurrent software, personnel, consumables, and support based on recall, again cross referencing our prepopulated spreadsheet and revising as necessary. All estimates were discussed collaboratively across staff during the meeting to generate the best consensus estimate.

We used these line items to calculate current costs. Additionally, healthcare facility staff were asked to describe the additional infrastructure, goods, and services that they needed to achieve and sustain WASH services. These items, and their associated quantities and unit prices were added to the spreadsheet to estimate the additional investment needed.

#### Stage 4 Sharing and discussion workshop

We held a two-day workshop to share and discuss the findings from preliminary costing. During the first day, we presented an overview of the costing spreadsheet and the preliminary data to the healthcare facility in-charges and the municipality health coordinator. Healthcare facilities agreed to conduct a second round of costing to improve the accuracy of preliminary costing and establish a system for routine costs monitoring. During the second day, the mayors and other municipality staff joined the meeting. We again presented an overview of the costing spreadsheet and preliminary data. We discussed possible elements of the O&M policy, given preliminary costs.

Municipal officials critiqued preliminary cost estimates as being too high and requested that healthcare facilities formally certify the data. We refined the costing spreadsheet to allow for additional space for healthcare facilities to provide comments as a form of budget justification. We finalized the costing spreadsheet with approval from municipal government and the project committee, who endorsed a second round of “formal” data collection where healthcare facility leaders were asked to officially certify the accuracy of the data.

#### Stage 5 Data validation and certification

We revisited all healthcare facilities and again explained the purpose of costing. This round, a broader range of staff were included in the meetings to triangulate the accuracy of data. Healthcare facility in-charges identified and delegated knowledgeable staff to review and revise costs as necessary, using the same bottom-up costing process. After data were collected, the project team conducted a second visit to present and review the data, at which time the healthcare facility staff verbally endorsed its accuracy. Healthcare facility In-charges then provided the final data and a signed letter certifying its accuracy.

#### Stage 6 Budget calculations

Using validated spreadsheets, we estimated current costs and costs of additional upgrades necessary to reach basic service. A small number of essential line items for capital hardware were missing from the certified data for some facilities, notably for drainage, waste processing areas, autoclaves, and/or fencing. In these cases, we imputed using the mean cost from other facilities. The final spreadsheets with all line items for each facility are included in Supplemental Files 1-8, and imputed data are notated for transparency.

We annualized capital hardware and capital software costs as the equivalent annual costs, using the time period as the estimated lifespan of the infrastructure in years (determined in consultation with the healthcare facility by an engineer from Geruwa, the local NGO supporting the project) and an annual interest rate of eight percent.^15,16^ These expenses represented large one-time investments to achieve WASH services (e.g., installation of infrastructure and start-up trainings for O&M), which are often financed through loans and repaid in installments. The annualized cost approximates the annual repayment amount.

Costs for capital maintenance, recurrent training, personnel, consumables, and support were routine expenses paid out of healthcare facilities’ normal annual operating budgets. These were already estimated as annual costs and required no further calculation.

We collected data in Nepalese rupees. Appendices record costs in Nepali rupees. Costs reported in this paper are converted to United States dollars (1 USD = 130.5 NPR).

#### Stage 7 O&M Policy finalization and approval

The policy formulation committee drafted an O&M policy for WASH in healthcare facilities based on the following information provided by the task force: (1) discussions with healthcare facility users as “rights holders” entitled to safe healthcare, civil society organizations, and duty bearers for ensuring adequate WASH (i.e., healthcare facility staff and management committees), (2) discussion and suggestions from the workshop from the Sharing and Discussion Workshop in Stage 4, and (3) results of budget calculations in Stage 6. This information was used to create a draft policy, based on a standard template from the municipality.

The draft was then reviewed by members of the project team (i.e., representatives from the NGOs Terre des hommes and Geruwa) and the municipal WASH coordination committee, who provided feedback. The policy formulation committee incorporated this feedback into a final draft, which was submitted by the deputy mayor, on behalf of the committee, to the municipal assembly meeting. The municipal assembly, headed by the mayor, approved the policy.

#### Stage 8 Dissemination and advocacy

At the municipal level, the municipality conducted dissemination workshop to ensure that all healthcare facilities were aware of the new policy, including newly established funds for operations and maintenance. At the district level, we held a half-day dissemination and advocacy workshop with healthcare facility in-charges, representatives from other municipal governments, local NGOs, the district health authority, and representatives from the media. We provided an overview of the costing and policy development process to raise awareness and encourage replication in other municipalities throughout the district. Thakurbaba municipality organized a similar workshop at the province-level on July 30^th^ 2023, where the Chief Minister of the Lumbini Province was the chief guest. During this workshop, the municipality officials shared their best practices and learnings from O&M policy development and advocated for its replication in other municipalities in the province and allocations of funds for WASH in healthcare facilities O&M from the provincial government. Similar workshops are planned at the national-levels for dissemination, but—at the time this manuscript was written—have not yet occurred.

### Ethics

This study was classified as not human subjects research by the Institutional Review Board of the University of North Carolina at Chapel Hill. Study activities received permission from local authorities of Thakurbaba municipality and from the in-charges of each healthcare facility.

## Results

### Costs of basic service provision

Spreadsheets complete with all line items for each facility are in Supplemental Files 1-8. Table 3 indicates the upfront capital hardware and software investments needed to achieve basic service; Table 4 indicates annual O&M costs and estimates of annualized capital costs. Table 5 disaggregates annual costs by WASH service.

**Table 3.** Upfront capital investments required to install WASH services. Costs are reported in United States dollars.

|  | Neulapur<br>health<br>post | Bagnaha<br>health<br>post | Shivapur<br>health<br>post | Thakurdw<br>ara health<br>post | Ranipur<br>basic<br>health<br>center | Bankatti<br>basic<br>health<br>center | Mohanpur<br>basic<br>health<br>center | Thakurdw<br>ara basic<br>health<br>center |
| --- | --- | --- | --- | --- | --- | --- | --- | --- |
| Upfront investment costs |  |  |  |  |  |  |  |  |
| Capital hardware cost | \$43,893 | \$38,330 | \$38,347 | \$15,954 | \$24,629 | \$10,123 | \$21,379 | \$21,448 |
| Capital software | \$368 | \$368 | \$460 | \$1,149 | \$368 | \$368 | \$368 | \$1,149 |

**Table 4.** Annualized costs for capital investments and operations and maintenance of WASH in healthcare facilities in Thakurbaba municipality. Total annual capital costs include capital hardware and capital software. Total annual operations and maintenance costs include capital maintenance, recurrent training, consumables, personnel, support. Costs are reported in United States dollars.

|  | Neulapur<br>health<br>post | Bagnaha<br>health<br>post | Shivapur<br>health<br>post | Thakurdwar<br>a health post | Ranipur<br>basic<br>health<br>center | Bankatti<br>basic<br>health<br>center | Mohanpu<br>r basic<br>health<br>center | Thakurdwar<br>a basic<br>health center |
| --- | --- | --- | --- | --- | --- | --- | --- | --- |
| Annual number of outpatient visits | 17016 | 8,004 | 3,558 | 7,992 | 2,315 | 3,937 | 5,104 | 2,525 |
| **Current annual expenditure** |  |  |  |  |  |  |  |  |
| Capital hardware<sup>‡</sup> | \$4,341 | \$3,607 | \$3,723 | \$1,564 | \$503 | \$733 | \$984 | \$1,170 |
| Capital software<sup>‡</sup> | \$0 | \$0 | \$0 | \$117 | \$0 | \$0 | \$0 | \$117 |
| Capital maintenance | \$1,728 | \$1,498 | \$2,696 | \$2,018 | \$845 | \$1,368 | \$1,516 | \$1,596 |
| Recurrent training | \$0 | \$0 | \$0 | \$0 | \$0 | \$0 | \$0 | \$0 |
| Consumables | \$1,476 | \$920 | \$1,616 | \$1,158 | \$1,048 | \$670 | \$808 | \$922 |
| Personnel | \$0 | \$0 | \$0 | \$0 | \$747 | \$0 | \$0 | \$0 |
| Support | \$0 | \$0 | \$0 | \$0 | \$0 | \$0 | \$0 | \$0 |
| **Additional annual investment needed to reach basic service** |  |  |  |  |  |  |  |  |
| Capital hardware<sup>‡</sup> | \$102 | \$102 | \$84 | \$113 | \$2,470 | \$274 | \$1,089 | \$972 |
| Capital software<sup>‡</sup> | \$37 | \$37 | \$47 | \$0 | \$37 | \$37 | \$37 | \$0 |
| Capital maintenance | \$184 | \$184 | \$156 | \$153 | \$153 | \$184 | \$153 | \$153 |
| Recurrent training | \$755 | \$648 | \$252 | \$578 | \$578 | \$578 | \$578 | \$578 |
| Consumables | \$315 | \$193 | \$547 | \$529 | \$457 | \$447 | \$441 | \$838 |
| Personnel | \$274 | \$274 | \$403 | \$274 | \$274 | \$274 | \$274 | \$473 |
| Support | \$316 | \$316 | \$170 | \$363 | \$316 | \$316 | \$316 | \$162 |
| **Summary costs** |  |  |  |  |  |  |  |  |
| Total annual capital costs | \$4,480 | \$3,746 | \$3,854 | \$1,794 | \$3,010 | \$1,044 | \$2,110 | \$2,259 |
| Total annual operations and maintenance costs | \$5,047 | \$4,033 | \$5,841 | \$4,593 | \$4,417 | \$3,837 | \$4,086 | \$4,722 |
| Total annual cost for basic service | \$9,527 | \$7,779 | \$9,695 | \$6,387 | \$7,427 | \$4,881 | \$6,196 | \$6,981 |
| Costs per outpatient visit | \$0.56 | \$0.97 | \$2.72 | \$0.80 | \$3.21 | \$1.24 | \$1.21 | \$2.76 |
| <sup>‡</sup>Capital hardware and capital software costs are annualized from the total upfront investment cost, using an interest rate of 0.08 |  |  |  |  |  |  |  |  |

**Table 5.** Total annual costs for disaggregated by water, sanitation, hygiene, cleaning, and waste management. “Other” category includes fencing and costs that were shared across multiple categories (e.g., infection prevention training, operations and maintenance training common to all infrastructure). Costs are reported in United States dollars.

| WASH component | Neulapur<br>health post | Bagnaha<br>health post | Shivapur<br>health post | Thakurdwa<br>ra health<br>post | Ranipur<br>basic<br>health<br>center | Bankatti<br>basic<br>health<br>center | Mohanpur<br>basic<br>health<br>center | Thakurdwa<br>ra basic<br>health<br>center |
| --- | --- | --- | --- | --- | --- | --- | --- | --- |
| Water | \$1,017 | \$619 | \$1,068 | \$781 | \$654 | \$527 | \$577 | \$680 |
| Sanitation | \$2,589 | \$1,413 | \$1,660 | \$1,197 | \$1,040 | \$752 | \$1,123 | \$932 |
| Hygiene | \$226 | \$146 | \$375 | \$45 | \$253 | \$220 | \$296 | \$67 |
| Waste management | \$483 | \$348 | \$387 | \$277 | \$1,069 | \$160 | \$121 | \$100 |
| Cleaning | \$2,681 | \$1,788 | \$3,541 | \$2,324 | \$1,457 | \$1,582 | \$1,667 | \$2,225 |
| Other | \$2,532 | \$3,466 | \$2,664 | \$1,763 | \$2,955 | \$1,640 | \$2,412 | \$2,537 |

The annualized cost for all basic WASH services ranged from $4,881 to $9,527 per year ($0.56-2.76 per patient visit). We did not find a meaningful trend in costs by type of facility or patient volume. Larger facilities had lower costs per patient visit, simply due to the higher volume of patients treated. The largest single contributor to annual costs was capital hardware, of which the primary cost drivers were construction of infrastructure for water supply, sanitation, waste management, and fencing. However, the combined annual O&M costs (capital maintenance, recurrent training, consumables, personnel, and support) exceeded annualized capital costs in all facilities.

Additional investment was needed to achieve basic service for all cost categories. The areas of greatest need were consumables, recurrent training, and capital software. All facilities reported needing additional waste management (waste bags and containers) and cleaning (detergents, disinfectants, and tools such as mops and brooms) supplies to reach targets for basic service. No facility had current expenditure for recurrent training, and few had line items for capital software. All facilities identified additional line items for trainings on infection prevention and control, infrastructure O&M, and cleaning were needed to reach basic service. Few facilities had current expenditure on personnel and included line items for support staff and O&M focal persons as additional expenditures needed to reach basic service.

On average, the highest annual expenditure was for cleaning, then sanitation. Cleaning costs were driven primarily by consumables (e.g., detergents, mops, brooms) and personnel salaries to perform cleaning activities. Facilities with maternity services had particularly high cleaning costs. Sanitation costs were driven more by capital hardware and capital maintenance costs. The lowest costs were for hygiene and waste management, as these did not require substantial capital investments or consumables. Sinks for hand hygiene were relatively low cost, and waste was managed in most cases by low-cost burning.

### Policy outcomes

On March 25^th^ 2023, Thakurbaba municipal government formally adopted an O&M policy for WASH in healthcare facilities. The policy calls for the establishment of a recovery fund that can be used for WASH infrastructure repair and maintenance at any municipal healthcare facility. The amount allocated to the fund is not formally specified in the policy but has been agreed through meetings of the municipality general assembly as USD 3,831 (500,000 NPR), which has been allocated from the municipality’s discretionary funds. The fund does not have an expiration date, and the municipality aims to replenish the fund at the end of every fiscal year (or sooner if all funds are exhausted). The policy also aims to establish an additional recovery fund in each healthcare facility for minor repairs and maintenance of WASH infrastructure. The municipality aims to establish these funds at USD 383 (50,000 NPR), but to date only USD 153 (20,000 NPR) per healthcare facility has been allocated.

To draw from the recovery funds, healthcare facilities submit a procurement request using existing forms and processes. Expenditures of up to USD 38 (5000 NPR) can be made at the recommendation of the WASH FIT committee within each healthcare, or up to USD 766 (100,000 NPR) at the recommendation of the healthcare facility operations and maintenance committee. Expenditures more than this amount must be recommended and approved by the municipal WASH coordination committee. For minor O&M tasks, repairs may be handled in-house by cleaning and maintenance staff. For major O&M tasks, the fund can be deployed to hire private sector contractors.

The Chief Executive Officer of the municipality (or individual assigned by the them) is responsible for maintaining the records of decisions and expenditures from the municipal-level recovery fund. The municipality WASH coordination committee is responsible for monitoring spending and can recommend disciplinary actions if any misuse is identified. Individual healthcare facility recovery funds are jointly operated by the chairperson of the healthcare facility operation and maintenance and the healthcare facility in-charge.

The municipality also adopted a guideline called the “WASH in healthcare facilities O&M fund implementation procedure,” which outlines expected costs of WASH infrastructure and O&M (including necessary tools, parts, supplies, and personnel) and describes procedures for implementing O&M activities. At present, there are no specific targets for infrastructure coverage or functionality, though targets may be established in future based on ongoing assessments and development of national guidelines.

### National advocacy outcomes

At the national level, we shared an interim report on costing from this project with the team at the Ministry of Health and Population that is preparing the national “roadmap for water sanitation and hygiene facilities for Nepal.” The costing report has been included in the references as evidence to inform the national roadmap. A draft national roadmap has been shared with stakeholders for consultation, and—at the time of writing this manuscript—was nearing completion.

## Discussion

We conducted costing and advocacy activities in Thakurbaba municipality, Nepal, with the aim of developing budgets and an O&M policy for WASH in healthcare facilities. Through a joint exercise with concerned health system and municipal government actors, we successfully collected costs data in eight healthcare facilities, which were used to draft budgets for annual costs of WASH service provision. These budgets informed an O&M policy, which was formally adopted by the Thakurbaba municipal government. The municipal government has established a recovery fund of USD 3,831 to implement the policy and is preparing to advocate the adoption of such policy at the national level. The calculated costs of WASH provision have also been used to inform the nationally costed roadmaps for WASH in healthcare facilities.

While there have been previous studies costing WASH in healthcare facilities (see, e.g.^14,17–19^), to our knowledge this is the first study to document the application of costs data for improved policy and practice. Even where cost evidence is being generated, its translation into meaningful improvements in policy and practice has been slow. Collecting costs data in isolation is of limited value if those data cannot be translated into improved policy or practice. To-date, only 16% of countries reporting data for progress on the Eight Practical Steps have developed national coordination mechanisms and costed roadmaps.^3^

Prior studies have suggested four factors that can improve the translation of evidence into policy and practice: salience, credibility, legitimacy and timeliness.^20–22^ Salience refers to evidence that is highly relevant to critical health issues within the context. Credibility refers to the rigor or scientific credibility of the evidence. Legitimacy refers to the process of producing the information, and whether it has been balanced, fair, and respectful of stakeholders’ values. Timeliness refers to the time between generating evidence and disseminating to the relevant policy makers. Below, we reflect on lessons learned during our project that demonstrate these factors.

### Adapt costing tools to reflect local needs and standards (salience)

During our pilot testing, we assessed the resources that were used locally by healthcare facilities to provide basic WASH. While we referenced the JMP service levels, we also incorporated for infrastructure, goods, and services that healthcare facilities identified as locally relevant needs. This helped ensure that final budgets reflected salient WASH needs in the local context. As part of this project, we also established a monitoring system (described below), to understand typical functionality of WASH infrastructure in the Thakurbaba context. This information will be used to set targets that balance considerations of what is practical and achievable in the local context versus international guidelines and standards. We did not reference national or subnational guidelines (e.g., local ordinances or building codes), but these may also be relevant information sources in other settings.

### Include a variety of stakeholders in costing (credibility)

During costing, we incorporated a variety of staff in meetings to estimate costs. Healthcare facility staff were knowledgeable about costs of O&M (capital maintenance, recurrent training, personnel, and support), but struggled to estimate capital costs. We invited an engineer to participate in costing, who was more knowledgeable on construction costs for capital hardware. This reflects experiences with costing in other settings, which have found that knowledge of costs is highly compartmentalized and that including staff from different roles to improve accuracy.^12–14^ Triangulating results between different perspectives is also a well-accepted practice to improve data quality in many research disciplines.^23,24^ Healthcare facility in-charges or other senior leadership staff can help identify knowledgeable individuals to participate in and triangulate data collection.

### Create a data formal certification process (legitimacy and credibility)

We presented the results of preliminary data collection in a sharing and discussion workshop. During this workshop, municipal government officials critiqued the accuracy (i.e., credibility of data). In response to this, we initiated a second round of data collection to address these concerns (i.e., Stage 5: Data validation and certification). To address credibility concerns, the municipal government asked healthcare facilities to officially certify the data in a signed letter. By creating a formal certification process, this created pressure on healthcare facilities and compelled a wider range of knowledgeable staff to participate in data collection and verify its accuracy.

Certification also improved legitimacy. The interim sharing and discussion workshop allowed stakeholders to voice their concerns about the data collection process and results, and we modified our approach and incorporated a step for healthcare facilities to formally certify the data before it was used for budgeting and policy development. This certification process addressed stakeholder concerns and helped improve the legitimacy of the data. On our context, healthcare facilities certified the data by providing a signed letter attesting to its accuracy. However, in other contexts different mechanisms may be more appropriate to improve credibility and legitimacy. Workshops with policy makers and end users of the data can identify locally appropriate alternatives in other contexts.

### Engage policy makers in evidence generation (legitimacy)

Early in the project, the municipality created a policy formulation committee chaired by the vice mayor, and a task force to support the committee. This policy formulation committee did not participate in day-to-day activities for data collection but was engaged in sharing and dissemination workshops to receive updates after major data collection milestones. Forming the committee was important to show the municipal government’s support and approval for the project and endorse data collection activities. Keeping municipal government stakeholders informed via the task force was also important for transparency, building trust in the data, and ensuring that the municipality’s needs, concerns, and priorities were being addressed.

In Nepal, municipal government has strong autonomy over budgeting and policy making for WASH in healthcare facilities,^7^ and thus these were the key stakeholders we engaged on committees. However, in other countries, other stakeholders may be more relevant. Informal assessments or expert opinions of local project personnel before activities begin may be necessary in other contexts to identify key gatekeepers, governing bodies, and other stakeholders for data collection.

### Establish systems for ongoing data collection (timeliness)

We collected data through asking healthcare facility staff to estimate costs. These estimates were based in part on recall of prior expenses. During interim sharing and discussion workshops, stakeholders suggested adapting the data collection tools to an online dashboard. The project team selected Kobo Toolbox connected to online Power BI (a data visualization software) as the preferred data collection platform because of its simplicity, the team’s prior knowledge, and its fitness for purpose. The online dashboard is being established and piloted in two HCFS. It will collect periodic data on the functionality of all the WASH facilities, preventive maintenance activities, repairs (e.g., to damaged infrastructure) and the cost and response time of the repair, and use of hand hygiene facilities. This database is intended to help develop appropriate strategies and targets for improving functionality and use and to reduce the cost on O&M. Delivering this information through an online dashboard will improve the availability and timeliness of information for decision makers.

## Limitations

We estimated costs using bottom-up costing. Healthcare facility staff estimated the quantity and unit prices of resources used in WASH provision based on available records and their best recall. While we triangulated estimated costs between different staff members and asked healthcare facilities to certify data to improve accuracy, we still found that certified data were missing a small number of key line items. We imputed missing data for capital hardware that were essential to reach the JMP basic service level (e.g., autoclaves). However, there are currently no comprehensive guidelines on other cost categories (e.g., quantity of consumable products like soap needed for adequate cleaning). Bottom-up costing is naturally prone to underestimates, as it relies on a comprehensive accounting of line items to generate accurate estimates. As such, the true costs of WASH service provision presented here may be underestimates. However, we reference prior studies of essential resources for WASH provision and imputed the most expensive missing line items for capital hardware, so we propose that any missing line items would have only a marginal effect on cost estimates.

## Conclusions

A supporting policy environment and adequate funding are essential to achieving and maintaining WASH in healthcare facilities, and developing nationally costed roadmaps is recommended by the WHO and UNICEF as part of the Eight Practical Steps to achieving universal access to WASH in healthcare facilities. While Nepal has made considerable progress on steps for assessing conditions and documenting the need, and setting targets, developing budgets and allocating funding for long-term O&M remains a challenge. We conducted costing and advocacy in Thakurbaba municipality, Nepal to accompany development of an operations and maintenance policy for O&M of WASH in healthcare facilities.

Our efforts resulted in successful development and adoption of an O&M policy for WASH in healthcare facilities by Thakurbaba municipality. At the time of publication, advocacy efforts to replicate this process in other municipalities, and findings on costs had been incorporated into a draft nationally costed roadmap. We propose that the process described in this paper can be used as an example to guide progress towards universal access to quality WASH services in healthcare facilities in other settings—particularly following the framework of the Eight Practical Steps.

## Data Availability

All data produced in the present study are available upon reasonable request to the authors

## Declarations

### Author contributions

Conceptualization: LKC, PB, MV, JB, DMA. Data curation and investigation: LKC, PB, RCB, RHJ, SK, HS. Project administration: LKC, PB, DG. Methodology, formal analysis, writing – original draft: LKC, PB, DMA. All authors reviewed and approved the final draft.

## Acknowledgements

We acknowledge Mr. Prem P. Giri from Terre des hommes Nepal and Ms. Nirmala Rija from Geruwa for their support with project management. We thank Ryan Cronk and Aaron Salzberg for their feedback on drafts of this manuscript. We also are extremely grateful to the healthcare facility staff and in-charges, municipality staff, and elected officials who participated in this project and contributed their valuable time and expertise.

## Disclaimer

N/A

